# Predictive Analysis for COVID-19 Spread in India by Adaptive Compartmental Model

**DOI:** 10.1101/2020.07.08.20148619

**Authors:** S. S. Singh, D. K. Mohapatra

**Affiliations:** Safety Research Institute, Atomic Energy Regulatory Board, Kalpakkam, 603 102

## Abstract

The role of mathematical modelling in predicting spread of an epidemic is of vital importance. The purpose of present study is to develop and apply a computational tool for predicting evolution of different epidemiological variables for COVID-19 in India. We propose a dynamic SIRD (Susceptible-Infected-Recovered-Dead) and SEIRD (Susceptible-Exposed-Infected-Recovered-Dead) model for this purpose. In the dynamic model, time dependent infection rate is assumed for estimating evolution of different variables of the model. Parameter estimation of the model is the first step of the analysis which is performed by least square optimization of priori data. In the second step of the analysis, simulation is carried out by using evaluated parameters for prediction of the outbreak. The computational model has been validated against real data for COVID-19 outbreak in Italy. Time to reach peak, peak infected cases and total reported cases were compared with actual data and found to be in very good agreement. Next the model is applied for the case of India and various Indian states to predict different epidemiological parameters. Priori data was taken from the beginning of nation-wide lockdown on 24 March to 6 July. It was found that peak of the outbreak may reach in the month of August-September with maximum 4-5 lakhs active cases at peak. Total number of reported cases all over India would be in between three to five millions. State wise, Maharashtra, Tamilnadu and Delhi would be worst affected.

## 1. Introduction

COVID-19 epidemic originated from Wuhan, the capital of China’s Hubei province in December 2019 [1]. Since then the epidemic has spread to all over the world. Considering the scale and speed of transmission of COVID-19, on 11 March 2020, the World Health Organization (WHO) declared it as a pandemic [2]. It has shown rapid infections in almost all countries. Till date, there is no specific vaccine, antivirals or effective therapeutics to treat coronavirus infections. As of 7 July 2020, a total 11,954,944 cases are confirmed all over the world. There are 4,505,864 active cases and 546,720 deaths [3].

The first COVID-19 case in India was reported in 30 January 2020 in the state of Kerala. As of 7 July 2020, there are 264,944 active cases, 456,830 discharged cases and 20,642 deaths in all over India [4]. Six cities i.e. Mumbai, Delhi, Ahmedabad, Chennai, Pune and Kolkata account for around half of the reported cases in the country. On 22 March 2020, India observed a 14-hour voluntary public curfew followed by a nationwide lockdown since March 24, 2020. Several other measures such as quarantine of the suspected cases, public health guidelines on social distancing, frequent hand washing and wearing face mask while stepping out of home for essential service are enforced.

Modelling of time evolution of an epidemic is essential for short-term and long-term planning for managing the outbreak. Several kinds of models have been proposed for describing the time evolution of epidemics, which can be distinguished in two main groups: collective models and networked models. Collective models are characterized by a small number of parameters and describe the epidemic spread in a population using a limited number of collective variables. They include generalized growth models [5], logistic models [6], Richards models [7], Generalized Richards models [5], sub epidemics wave models [8], Susceptible-Infected-Recovered (SIR) models [6], [9], and Susceptible-Exposed-Infectious-Removed (SEIR) models [5]. SIR, SEIR and other similar models belong to the class of the so-called compartmental models [5], [10].

Several studies were carried out to model the outbreak of COVID-19 in different countries. In their paper Poonia and Azad [11], applied Holt’s second order smoothing method and autoregressive integrated moving average (ARIMA) model to generate 10-day ahead forecast. Khajanchi et al. [12] studied an extended SEIR model to study the transmission dynamics of COVID-19 and perform a short-term prediction based on the data from India. A discrete-time SIR (Susceptible-Infectious-Removed) model introducing dead compartment system was studied by Anastassopoulou et al. [13]. Wu et al. [14] studied a SEIR model to investigate the dynamics of 2019-nCoV human-human transmission dynamics based on the data from Wuhan, China from December 31, 2019 to January 28, 2020. Wu et al. [15] used a SIR model to delineate the transmission dynamics of COVID-19 and also estimate the clinical severity for the coronavirus. To study the dynamics of COVID-19, a stochastic transmission model was also developed by Kucharski et al. [16].Fanelliet. al. used SIR (susceptible-infectious-removed) model introducing dead compartment system to analyse and forecast COVID-19 spreading in China, Italy and France.[17].Calafioreet. al. described a variant of the SIR model to describe the actual number of infected individuals[18]. Piccolominiet. al. proposed a modification of SEIRD by introducing a time dependent transmitting rate[19].

In this work, a variant of the SIRD and SEIRD model is proposed. A dynamic SIRD and SEIRD model have been chosen as the basic mathematical models. The infection rate is assumed to decrease exponentially with time as a result of various mitigation measures enforced by the government. As a first step of the analysis, the model parameters are estimated from the actual data by optimization of the model with actual data. The evaluated parameters are then used in simulation for prediction. A computer program has been developed indigenously for this purpose. The computational scheme has been validated against Italian data. Finally, the tool is used for India specific data to predict the COVID-19 evolution. In section 2 the details of both SIRD and SEIRD models are described. In section 3 validation of the computational tool is explained for Italian data. In section 4, India specific analysis has been described.

## 2. Numerical Model and Methods

In the present study, time dependent SIRD model has been employed. In this model the population N is divided into sub-population of susceptible (S), Infected (I), recovered (R) and dead (D) for all times t. Thus, N = S + I + R + D. The mean-field kinetics of the SIRD epidemic evolution is described by the following system of differential equations

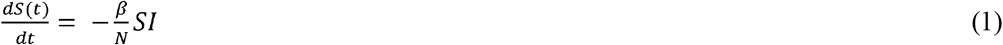

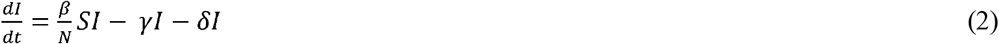

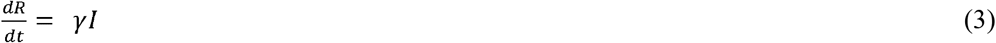

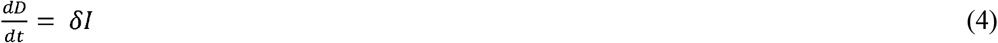

with initial condition [S(t_0_),I(t_0_), R(t_0_), D(t_0_)] = [S_0_,I_0_,R_0_,D_0_] for some initial time *t*_*0*_. The parameter β is the infection rate, i.e. the probability per unit time that a susceptible individual contract the disease when entering in contact with an infected person. The parameter γ and δ denote the recovery and death rates respectively. We have considered a modified version of the SIRD model, where the infection rate β is let vary with time. More precisely, given that the containment measures became law at time t^*^, we take

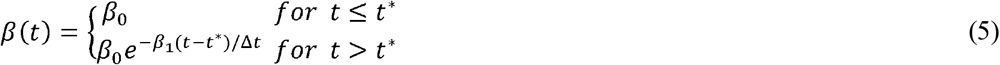

where β_0_ is the infection rate before start of the lockdown. t^*^ is the time at which lockdown has begun.

A more refined model is the SEIRD model where a new compartment E representing the exposed individuals that are in incubation period is added. The resulting equations in the SEIR model are the following:

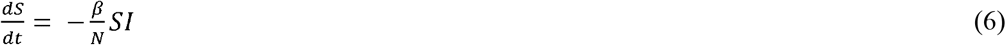

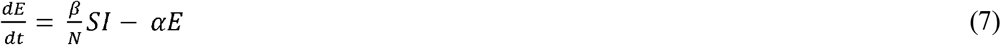

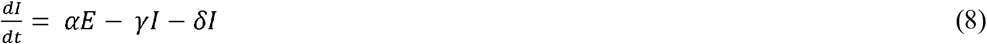

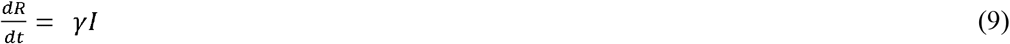

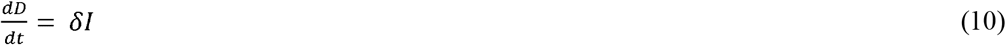

where α represents the incubation rate. The difference between the exposed (E) and infected (I) is that the former have contracted the disease but are not infectious and the later can spread the disease.

### 2.1 Dynamical model

In the present study, each dynamical model presented is either of the SIRD or SEIRD type. The SIRD and SEIRD simulations were implemented by the odeint function in Python’s SciPy library [20].

### 2.2 Parameter Estimation

The input required for the model is time series data of prior date and the population size. A standard dataset for COVID-19 consists of daily number of confirmed cases, recovered cases and deceased cases. Number of active cases is derived from the available data by subtracting the recovered cases and deceased cases from the confirmed cases. The variables in the model are susceptible population, exposed population, active population, recovered population and the deceased. From the time series data, the unknown parameters of the model e.g. S_0_, β_0_, β_1_, α, γ, and δ are determined by least square optimization. The system of ordinary differential equations is optimized with respect to the observation data. Objective function for the optimization is defined as chi-square which is given in Equation 11.

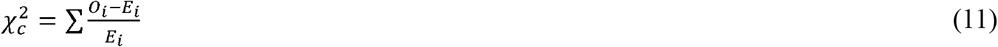

Where *O*_*i*_ is the observation and *E*_*i*_ is the modelled/expected value for a given instant. LMFIT module of Python library is used for the nonlinear least-square optimization [21].

### 2.3 Prediction of the epidemic outbreak

The evolution of the variables in the model is calculated by solving the set of differential equation by using the evaluated parameters from known information.

### 2.4 Limitations of the Predictive Model

The predictive tool in the present study uses a classical modified SIR model by deterministic method. The model is valid for a homogeneous population with no migration. In few instances it has been observed that initial conditions for the system of ordinary differential equation are zero. The optimization algorithm fails in such cases and the estimated parameters are same as the initial guess values. In some other cases there has been second surge in number of infected cases. Here also the optimization algorithm gives unphysical results. However for cases of India and most affected states the performance of the predictive tool has been quite satisfactory.

## 3. Validation against Italian Data

It is important to validate the model with actual data before applying for prediction of an epidemics outbreak. For this reason, an attempt has been made to validate both the predictive models against Italian data for which peak of COVID-19 epidemic has been reached.

For validation of the model, Italian time series data from date 22 February 2020 to 03 June 2020were taken from John Hopkins University’s data repository on COVID-19 [3]. The dataset contains cumulative confirmed cases, recovered cases and death cases for every day. In case of Italian data, peak of the epidemics was observed on 19 April 2020 with 108, 237 infected individuals. As on 03 June 2020, there were 236,142 total confirmed cases.

By using the developed model, validation exercise was carried out by taking first 40 days priori data for parameter estimation and then predicting evolution of model variables for remaining of the days. It has been observed that the prediction outcome is highly dependent on number of priori dataset for parameter estimation. Both SIRD and SEIRD models are used to check their validity as a predictive tool. Key results of the validation exercise are given in Table 1. The evolutions of active and confirmed cases are compared in Table 2. It is found that both models match reasonably well with the observation data. Prediction of number of active cases and total cases for case of Italy by 40 days priori data by using SEIRD and SIRD model are shown in Fig. 1 (a) and (b).

**Table 1:**
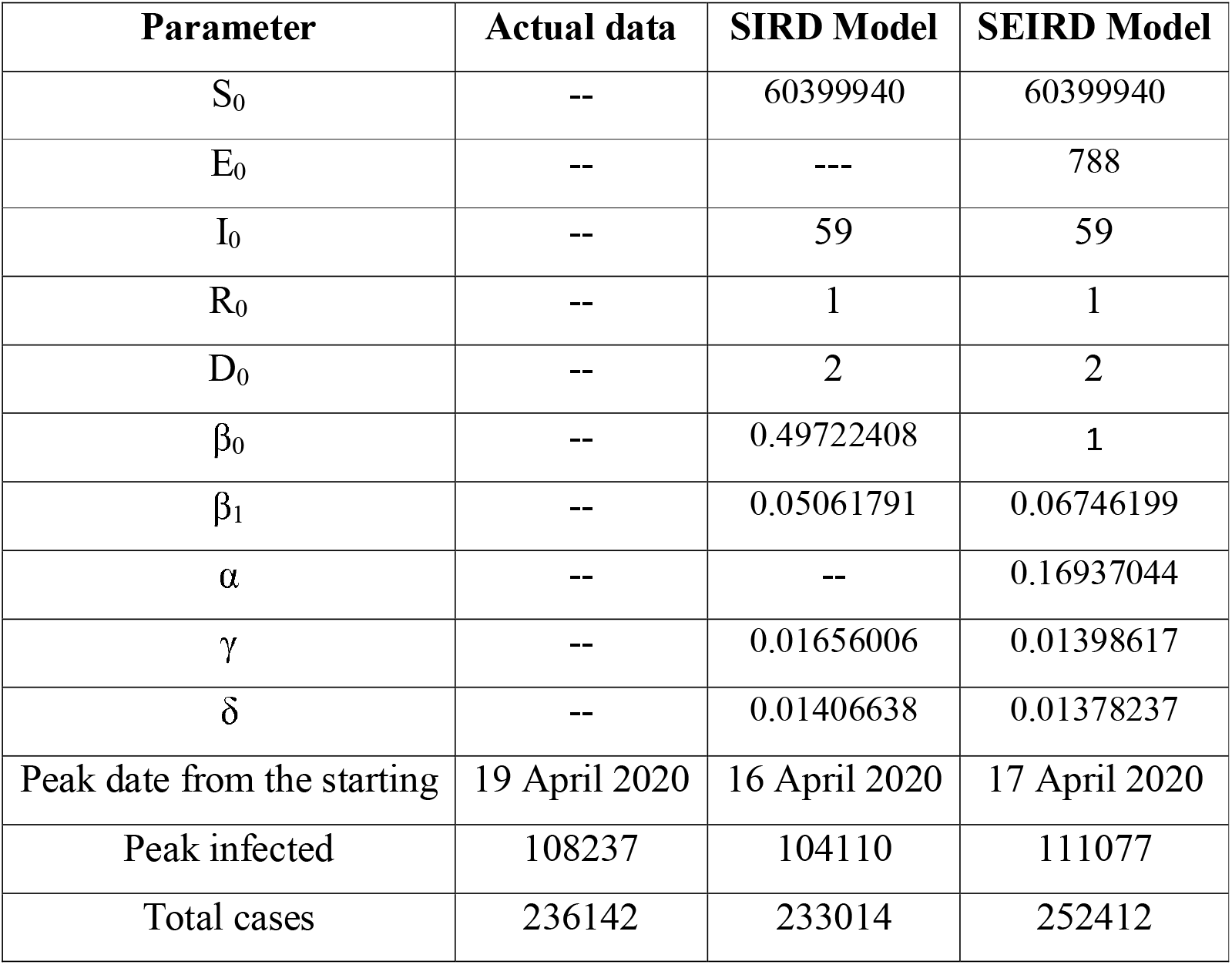
Key results of the validation exercise against Italian data.

**Table 2:** Comparison of the confirmed cases and active cases for the Italian data.

| Days | Confirmed Cases |  |  |  |  | Active cases |  |  |  |  |
| --- | --- | --- | --- | --- | --- | --- | --- | --- | --- | --- |
|  | Actual data | SIRD | Diff. | SEIRD | Diff. | Actual data | SIRD | Diff. | SEIRD | Diff. |
| 0 | 62 | 62 | 0 | 62 | 0 | 59 | 59 | 0 | 59 | 0 |
| 7 | 1128 | 966 | 162 | 738 | 390 | 1053 | 891 | 162 | 687 | 366 |
| 14 | 5883 | 6274 | -391 | 5814 | 69 | 5061 | 5610 | -549 | 5259 | -198 |
| 21 | 21157 | 22253 | -1096 | 22054 | -897 | 17750 | 19079 | -1329 | 19090 | -1340 |
| 28 | 53578 | 51878 | 1700 | 52084 | 1494 | 42681 | 42100 | 581 | 42499 | 182 |
| 35 | 92472 | 90631 | 1841 | 90665 | 1807 | 70065 | 68546 | 1519 | 68590 | 1475 |
| 42 | 124632 | 130120 | -5488 | 129777 | -5145 | 88274 | 90121 | -1847 | 89423 | -1149 |
| 49 | 152271 | 163807 | -11536 | 163874 | -11603 | 100269 | 101939 | -1670 | 101007 | -738 |
| 56 | 175925 | 189135 | -13210 | 190939 | -15014 | 107771 | 103706 | 4065 | 103439 | 4332 |
| 63 | 195351 | 206547 | -11196 | 211238 | -15887 | 105847 | 97891 | 7956 | 98917 | 6930 |
| 70 | 209328 | 217778 | -8450 | 225965 | -16637 | 100704 | 87622 | 13082 | 90061 | 10643 |
| 77 | 218268 | 224702 | -6434 | 236447 | -18179 | 84842 | 75520 | 9322 | 79084 | 5758 |
| 84 | 224760 | 228835 | -4075 | 243831 | -19071 | 70187 | 63347 | 6840 | 67562 | 2625 |
| 91 | 229327 | 231244 | -1917 | 249003 | -19676 | 57752 | 52102 | 5650 | 56493 | 1259 |
| 98 | 232664 | 232624 | 40 | 252615 | -19951 | 43691 | 42242 | 1449 | 46432 | -2741 |

**Fig 1:**
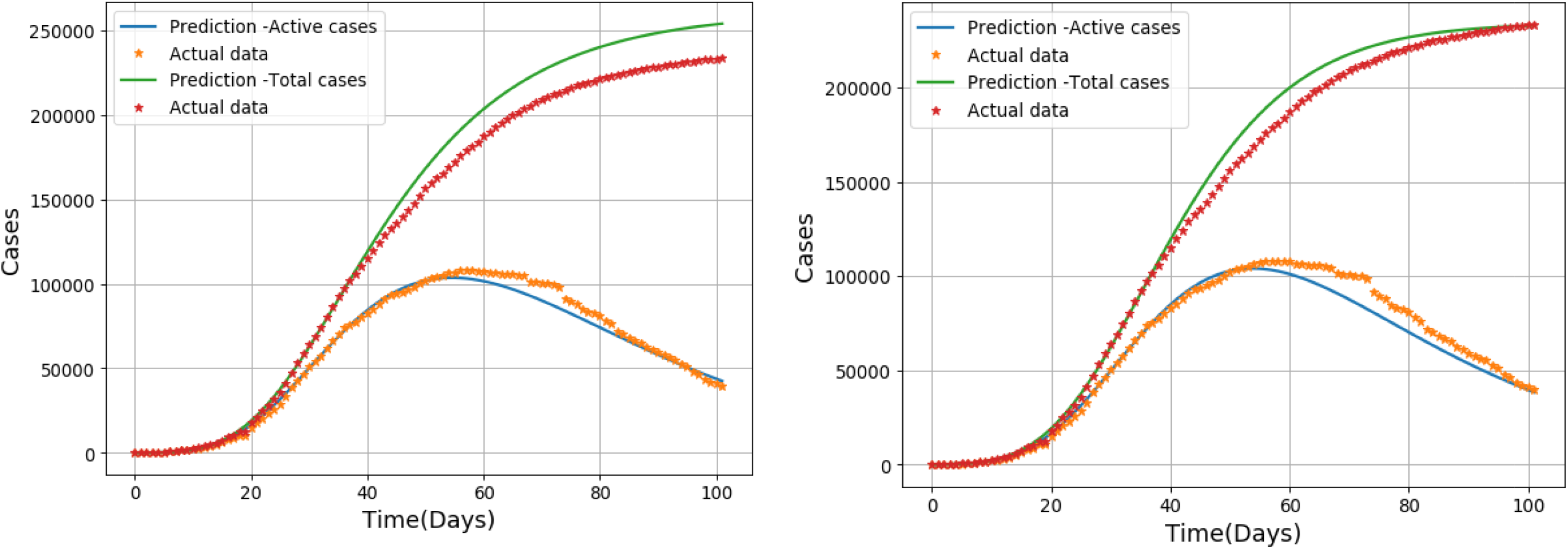
(a) and (b): Prediction of number of active cases and total cases for case for Italy by 40 days data by using SEIRD and SIRD model.

Infection rate at the beginning is estimated to be 0.49 and 1 per day by SIRD and SEIRD models. At the end of the time period, infection rate became 0.002 and 0 per day respectively. Incubation rate for the SEIRD model was found to be 0.17 per day. Recovery and mortality rate were estimated as 0.016 per day and 0.014 per day. The date of peak in infected cases was found to be 16 and 17 April by both the models which is in very good agreement with the actual date of 19 April 2020. Similarly peak infected cases match within five percent of the actual value. However, in case of total number of confirmed cases SIRD model matches well but a difference of more than 10% has been observed for the result of SEIRD model. As both models match fairly well with the actual data, both models are considered for the predictive analysis for India specific study.

## 4. Prediction for India Specific Data

For prediction of COVID-19 outbreak in India, raw data consisting of confirmed cases, recovered cases and deceased cases has been taken from the period of 24 March 2020 to 6 July 2020 from www.covid19india.org [22]. The predictive analysis closely follows the pattern followed by Sahoo et. al[23]. As per the calculation methodology, parameters for SIRD and SEIRD model were first determined from the data by least square optimization. The analysis is broadly divided into two specific categories e.g. India specific study and state specific study. In state specific study, predictive analysis of few selected states with large number of COVID-19 cases has been undertaken. Key parameters pertaining to the India specific study are given in Table 3. The infection rate at the beginning of nation-wide lockdown was found to be 0.16 per day and 0.38 per day by respective models. By 6 July 2020, infection rate by both models has come down 0.07 per day and 0.12 per day respectively. Similarly the incubation rate is found to be 0.03 per day. The recovery rate and death are found to be 0.05 per day (0.06 by SEIRD) and 0.002 per day respectively. The initial exposed individuals were estimated to be 9926 by LMFIT optimization.

**Table 3:**
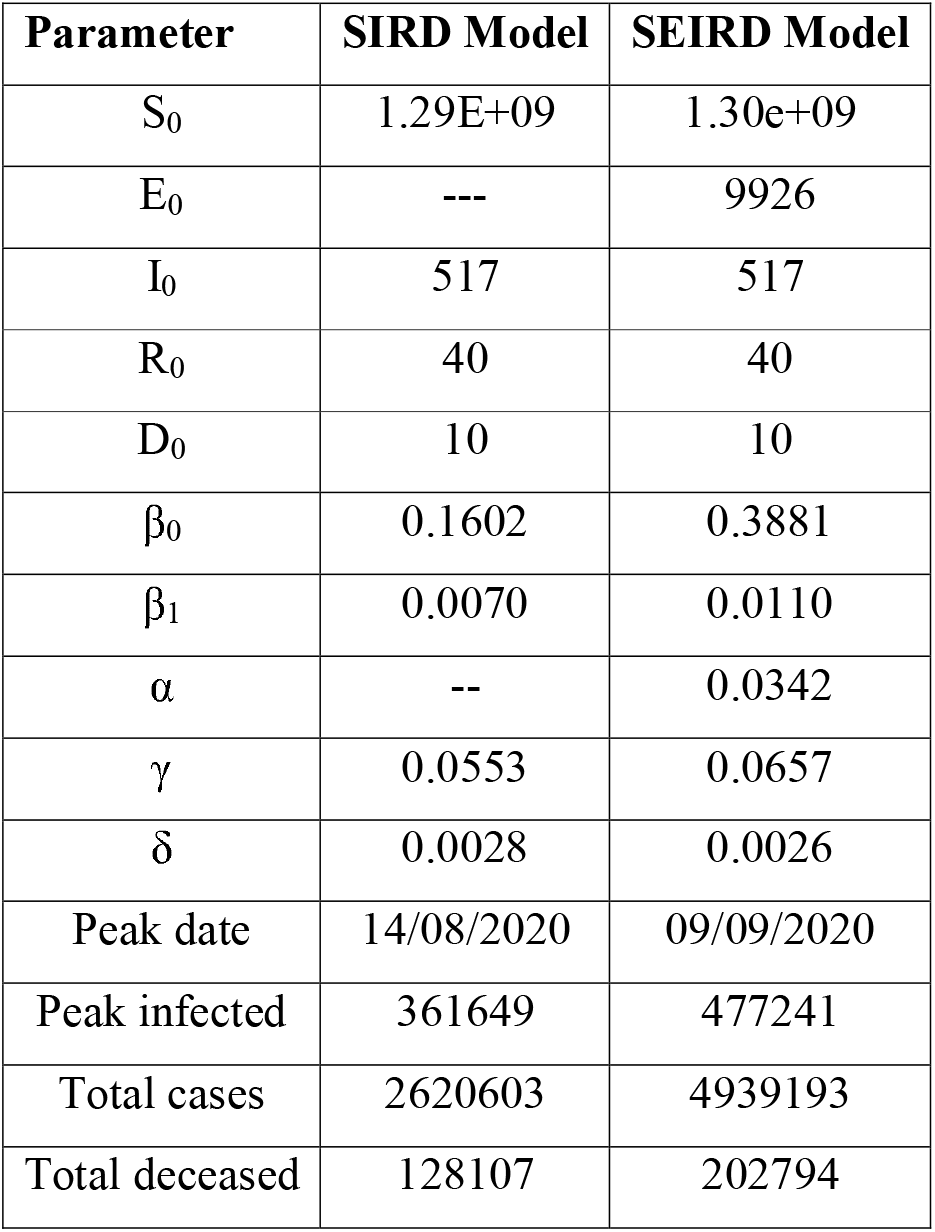
Key results for prediction in India.

Predictive analysis was carried out by taking the estimated parameters as input for the model. Simulation was carried out for a time period of 365 days starting from beginning of lockdown (24 March 2020) and evolution of all the SIRD variables were tracked. The evaluated parameters for both the models are given in Table 3. Evolution of SIRD and SEIRD variables are shown in Fig. 2 (a) and (b). It is found that by SIRD predictions the peak in infected individuals may reach in the second week of August. In case of the SEIRD model the same may occur in the first week of September. Similarly the peak infected would be 361K and 494K respectively. Total number of confirmed cases will be 2.6 million and 4.9 million respectively. Total number of casualties would be in between 100K and 200K. By January 2021, the epidemic would be almost over in India.

**Fig 2:**
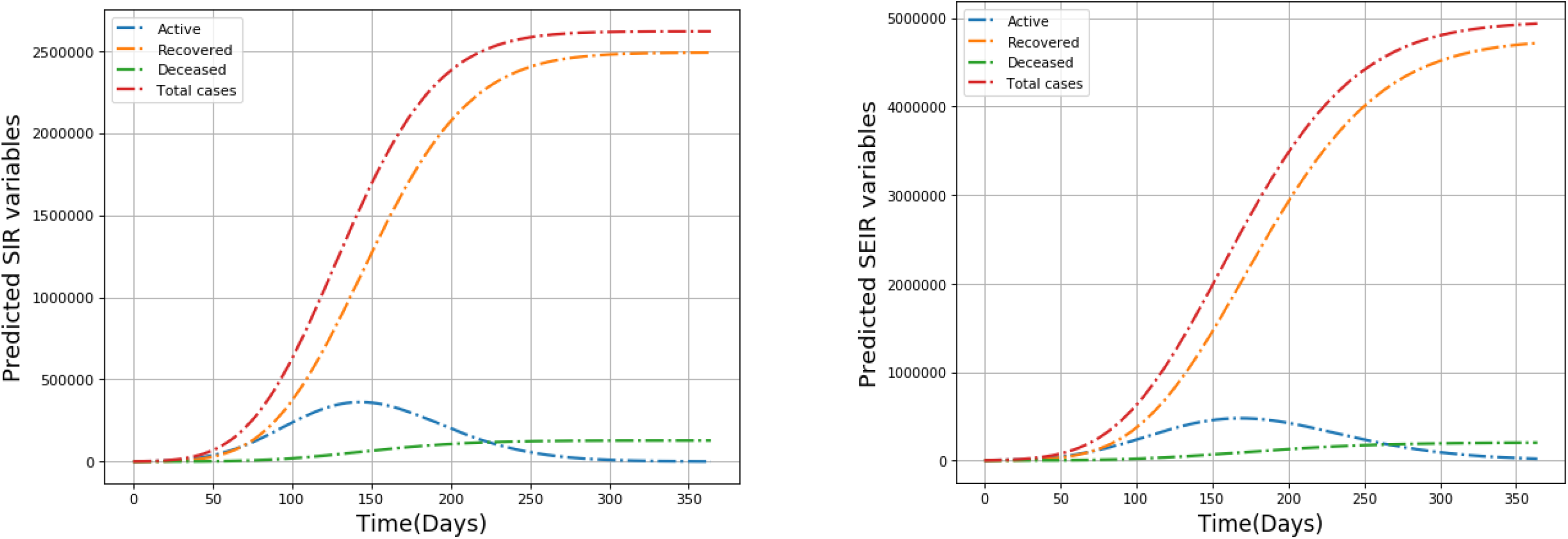
(a) and (b): Evolution of SIRD and SEIRD variables for predication in case of India.

### 4.1 Prediction of State Specific Results

Evolutions of total infected cases for different states by SEIRD model are shown in Fig. 3 (a) and (b). Similar results by SIRD model are shown in Fig. 4 (a) and (b). By both the models Mahrashra, Tamilnadu and Delhi may have around 500K predicted total infected cases during the epidemic. Gujarat, Madhyapradesh, Rajasthan, Uttarpradesh etc will be among the states which may have number of confirmed cases in between 10,000 to 100,000.

**Fig 3:**
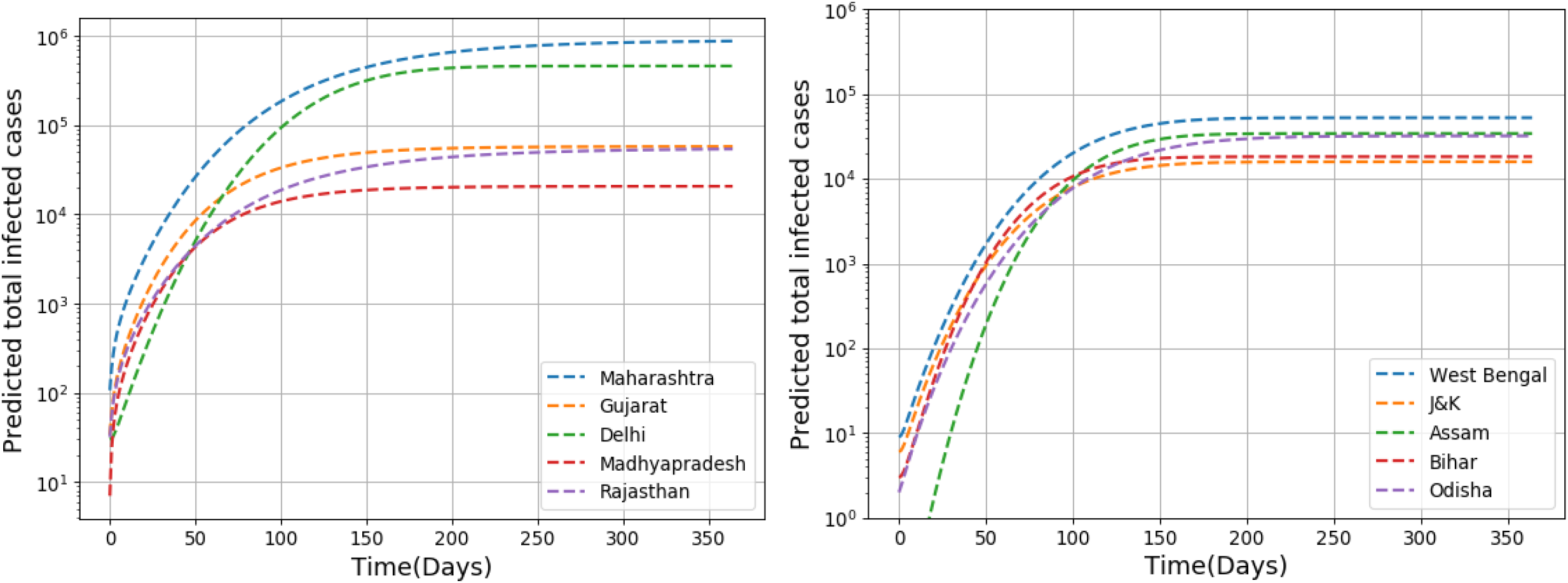
(a) and (b): Predicted total infected cases in various selected states of India by SEIRD model.

**Fig 4:**
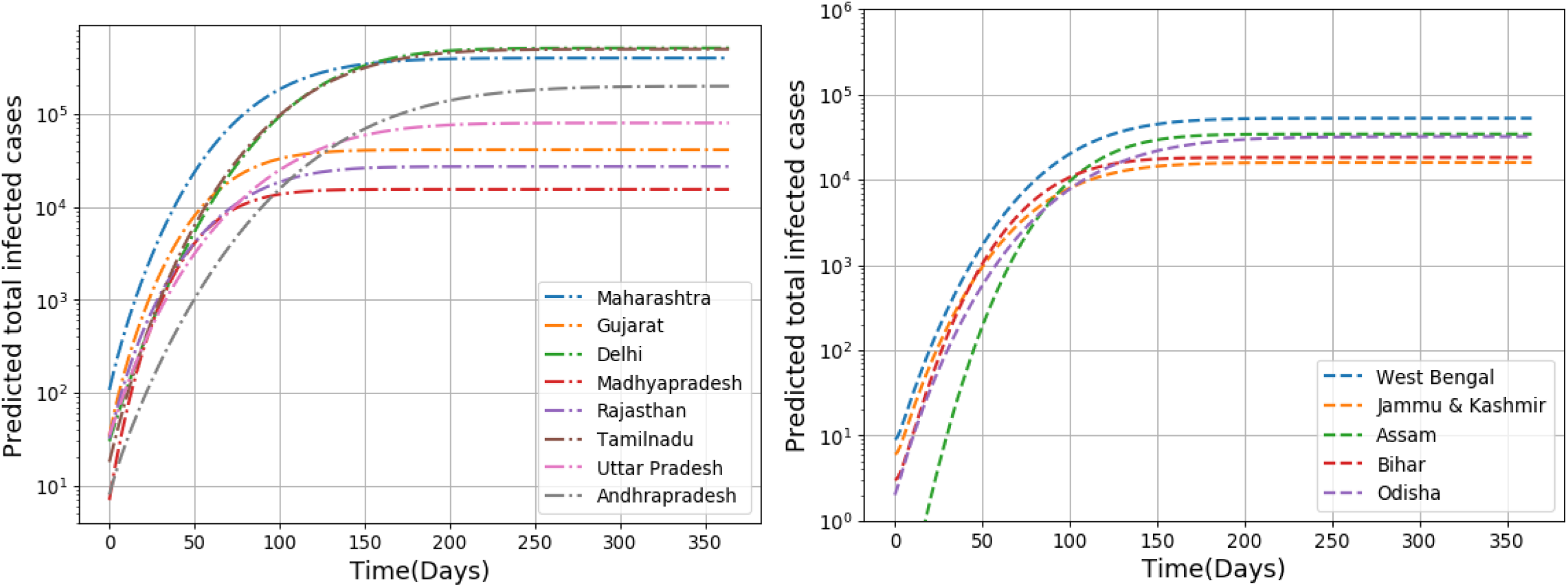
(a) and (b): Predicted total infected cases in various selected states of India by SIRD model.

Predicted new daily infected cases in various selected states of India by SEIRD model are shown in Fig. 5 (a) and (b). Similar results by SIRD model are shown in Fig. 6 (a) and (b). It is observed that in case of Maharshta daily new infected cases would be around five thousand in the month July. Similarly Tamilnadu and Delhi would experience more than five thousand cases in the months of July-August.

**Fig 5:**
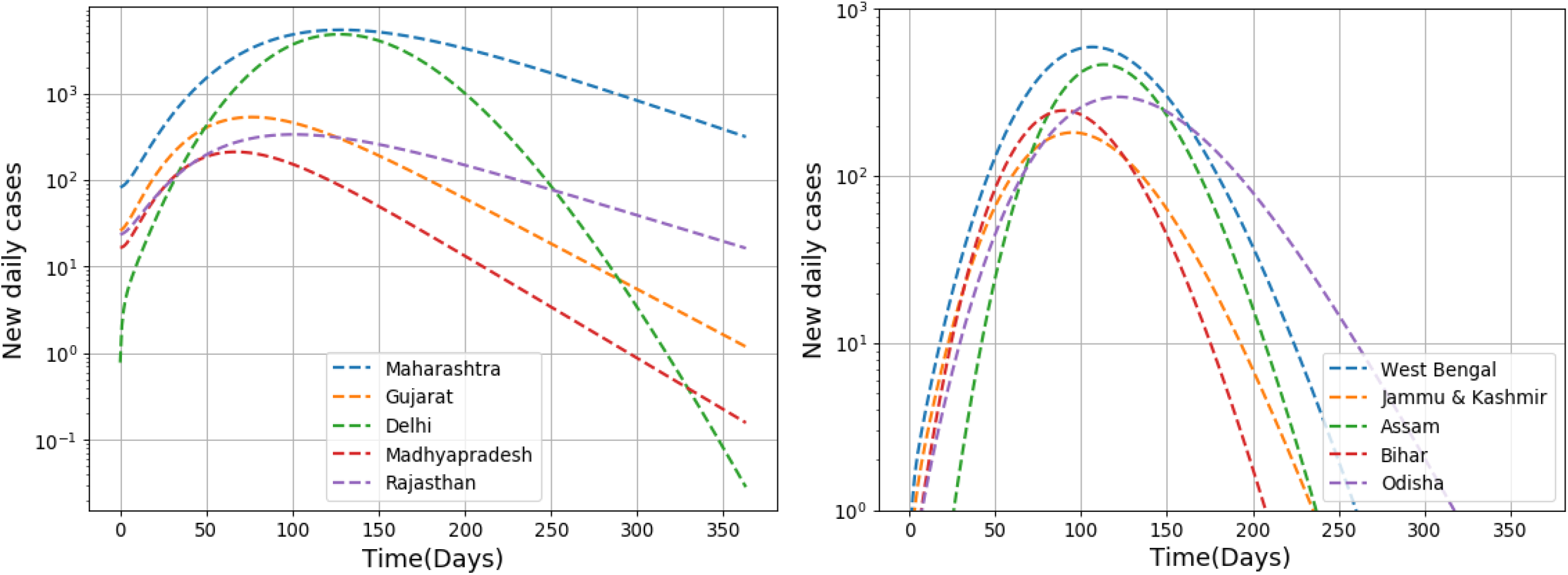
(a) and (b): Predicted new daily infected cases in various selected states of India by SEIRD model.

**Fig 6:**
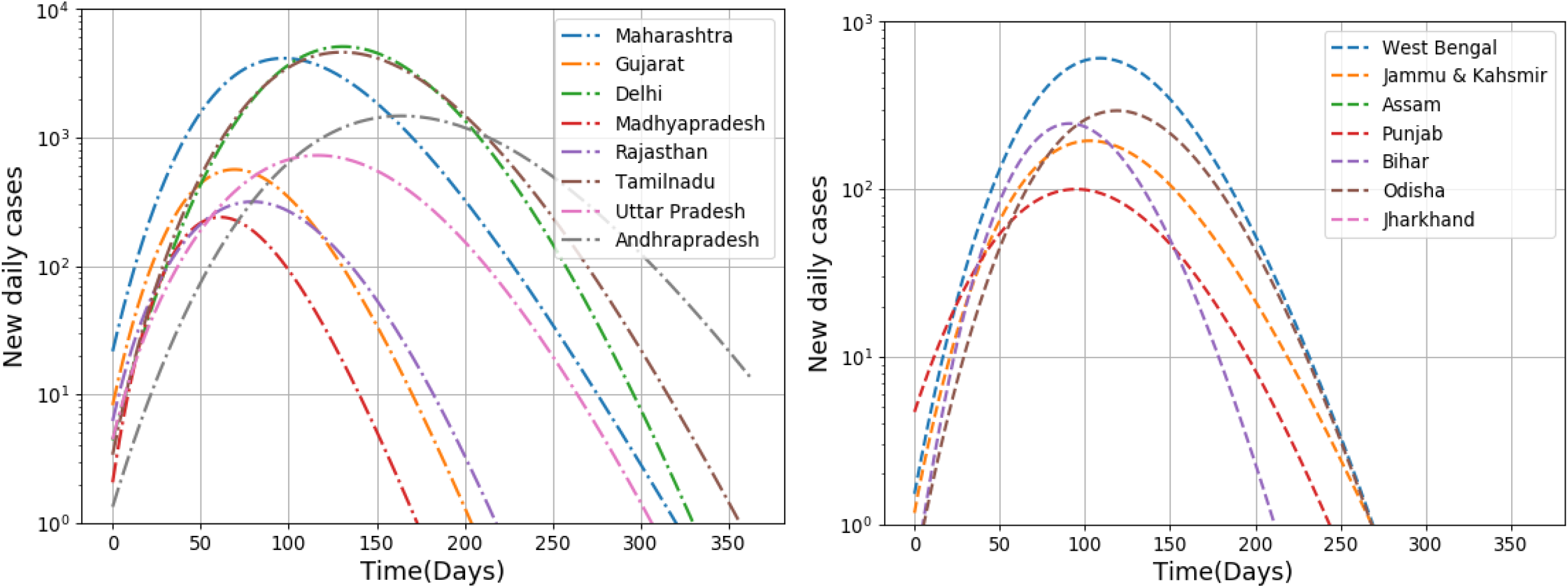
(a) and (b): Predicted new daily infected cases in various selected states of India by SIRD model.

Predicted active infected cases in various selected states of India by SEIRD model are shown in Fig. 7 (a) and (b). Similar results by SIRD model are shown in Fig. 8 (a) and (b). It is observed that the states of Maharashtra, Tamilnadu and Delhi would experiences active cases in between 10,000 to 100,000. Other states would experience less number of cases.

**Fig 7:**
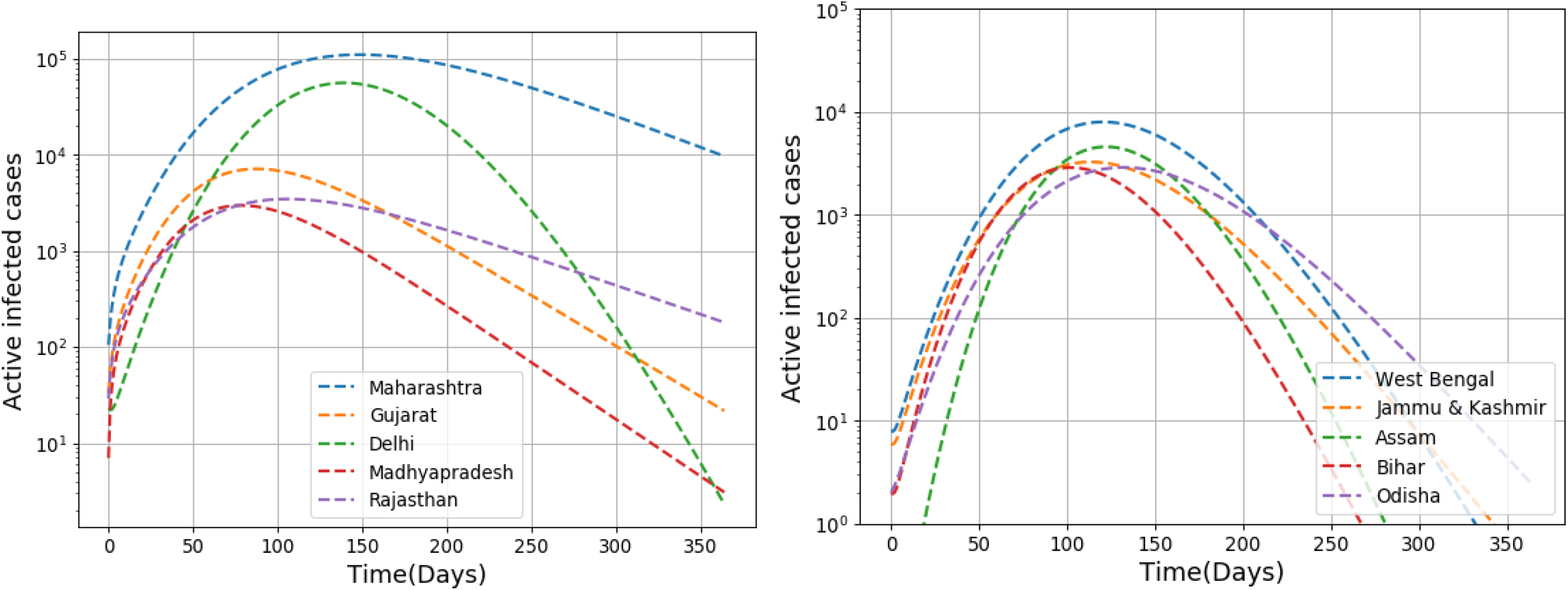
(a) and (b): Predicted active infected cases in various selected states of India by SEIRD model.

**Fig 8:**
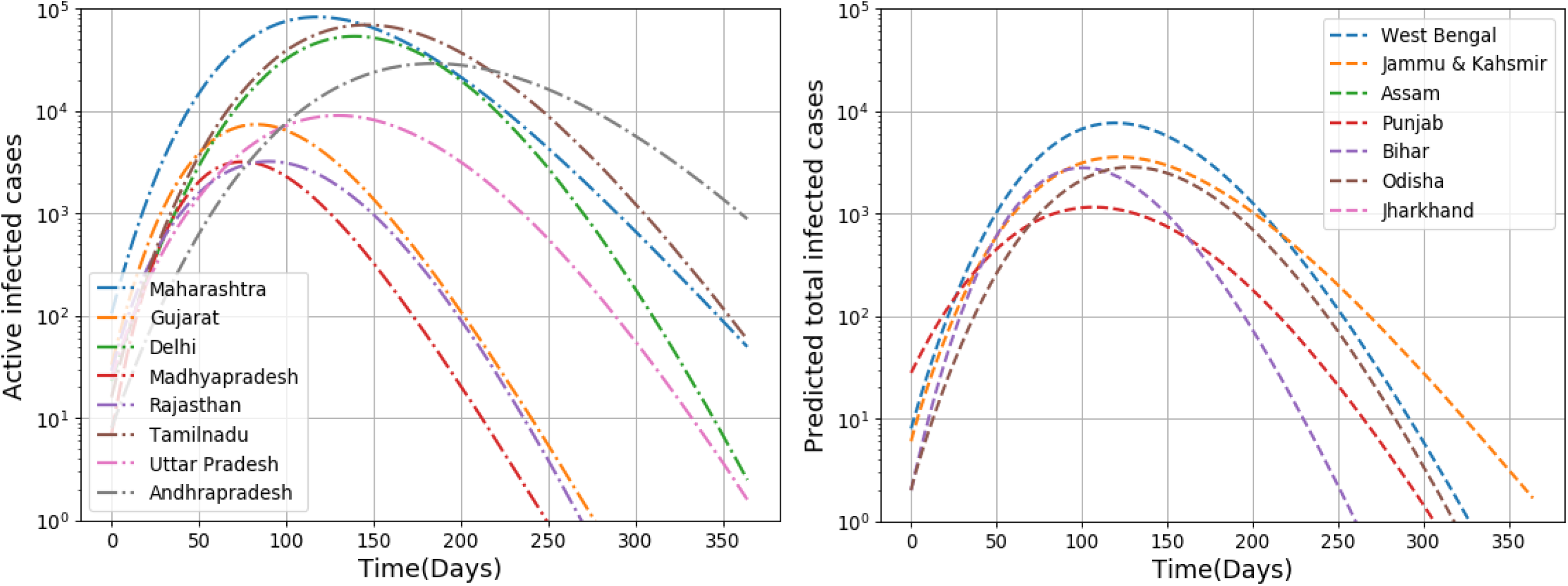
(a) and (b): Predicted active infected cases in various selected states of India by SIRD model.

### 4.2 Comparison of Prediction of peak and maximum infected cases

Table 4 compares state-wise results by SIRD and SEIRD models for the predicted time to reach peak of the epidemic, number of active, saturation of infected cases and Time to reach 99% of total infected cases. Results suggest that India as a whole could see the peak in August-September 2020. Punjab, Andhra Pradesh and Kerala have already seen the peak in the month of May followed by Gujarat, Uttar Pradesh, Rajasthan, Madhya Pradesh and Telengana in June 2020. Maharshtra, Tamilnadu and Delhi would see the peak in the month of July-August 2020.

**Table 4:** Comparison of results for various states of India.

| State | Time to reach peak (MM-YYYY) |  | Number of active cases at peak |  | Time to reach 99% of total infected cases (MM-YYYY) |  | Total expected infected cases |  |
| --- | --- | --- | --- | --- | --- | --- | --- | --- |
|  | SIRD | SEIRD | SIRD | SEIRD | SIRD | SEIRD | SIRD | SEIRD |
| India as a whole | Aug 2020 | Sep 2020 | 361,649 | 477,241 | Dec 2020 | Jan 2021 | 2,620,603 | 4,939,193 |
| Maharashtra | July 2020 | Aug 2020 | 83,320 | 109,708 | Nov 2020 | Feb 2021 | 397,550 | 880,717 |
| Gujarat | June 2020 | June 2020 | 7,411 | 7,107 | Sep 2020 | Dec 2020 | 41,013 | 57,855 |
| Delhi | Aug 2020 | Aug 2020 | 53,905 | 55,806 | Nov 2020 | Nov 2020 | 509,915 | 462,400 |
| Madhyapradesh | June 2020 | June 2020 | 3,195 | 2,963 | Aug 2020 | Nov 2020 | 15,402 | 20,711 |
| Rajasthan | June 2020 | July 2020 | 3,223 | 3,452 | Sep 2020 | Feb 2021 | 27,143 | 54,247 |
| Tamilnadu | Aug 2020 | --- | 69,838 | --- | Dec 2020 | --- | 496,725 | --- |
| Uttarpradesh | Aug 2020 | --- | 9,040 | --- | Nov 2020 | --- | 79,804 | --- |
| West Bengal | July 2020 | July 2020 | 7,710 | 7,989 | Oct 2020 | Oct 2020 | 56,054 | 52,697 |
| Jammu & Kashmir | July 2020 | July 2020 | 3,576 | 3,265 | Nov 2020 | Oct 2020 | 19,532 | 15,912 |
| Assam | --- | July 2020 | --- | 4,585 | --- | Oct 2020 | --- | 34,307 |
| Bihar | July 2020 | July 2020 | 2,809 | 2,880 | Sep 2020 | Sep 2020 | 18,756 | 18,276 |
| Odisha | Aug 2020 | Aug 2020 | 2,851 | 733 | Nov 2020 | Aug 2020 | 28,350 | 3,461 |

Number of active infected cases at peak for India would go up to 477 K by SEIRD model and 361 K in SIRD model. the most affected states-Maharashtra, Delhi and Tamil Nadu may see active infected cases up to 83K, 53K and 69K respectively. Uttar Pradesh, Rajasthan, Madhya Pradesh, Telengana, Odisha, Andhra Pradesh and Punjab may see active infected cases below 10K.

## 5. Conclusion

In the present work, a computational model for predicting spread of COVID-19 by dynamic SIRD and SEIRD model has been proposed. The dynamic model assumes time dependent infection rate. The model consists of estimating various model parameters and predicting the outbreak from estimated parameters. Predictive capability of the tool is highly dependent of number of priori data for parameter estimation. The tool has been validated against Italian data where the spread is evaluated from 40 days of data and compared with actual results. It is found that both SIRD and SEIRD model are in good agreement with the observational data. After validating, the tool is applied for prediction in India and different Indian states. Various epidemiological parameters such as time of peak arrival, number of active cases during peak, daily new cases and total number of reported cases are evaluated for the all cases. It is found that peak in India may arrive in the month of August-September with maximum number of active cases between 400-500 K. About 3-5 million reported cases will come during the outbreak. The epidemic spread will be over by the end of 2020. State wise, Maharashtra, Tamilnadu and Delhi would be most affected.

## Data Availability

Data are taken from public domain on COVID-19.

http://www.covid19india.org

